# Heritability of Age-Related Macular Degeneration in the Amish

**DOI:** 10.64898/2026.08.04.26359695

**Authors:** Noel C. Moore, Yeunjoo E Song, Alexander V. Gulyayev, Kristy Miskimen, Penelope Miron, Renee A Laux, Audrey Lynn, Sarada L Fuzzell, Sherri D Hochstetler, Dawn Miller, Laura J Caywood, Jason E Clouse, Sharlene D Herington, Ping Wang, Yining Liu, Daniel A Dorfsman, Jeffery M Vance, Muneeswar Gupta Nittala, SriniVas R Sadda, Dwight Stambolian, William K Scott, Margaret A Pericak-Vance, Jonathan L Haines

## Abstract

**Purpose:** Age-related Macular Degeneration (AMD), a degenerative disease of aging, leads to central vision loss and has a strong genetic risk. Genetic heritability, used to quantify genetic influence on a trait, has mainly focused on twin study designs but these are vulnerable to bias. Studying relatives beyond twins is necessary to bring clarity to the genetic burden of AMD and help focus the search for additional genetic risk loci.

**Methods:** Through both single nucleotide polymorphism (SNP) and pedigree-based heritability methods, the heritability of AMD was analyzed using relationship informed analyses of families from an Amish population (n = 525). AMD status was determined using the Beckman grading scale (285 controls and 240 cases). An estimate of genetic relatedness preceded SNP heritability estimation, whereas the pedigree heritability model utilized genealogical reports. Primary models were adjusted for age, sex, and population structure. A comparison of SNP- and pedigree-based models followed heritability estimation. Sensitivity models adjusting for all possible combinations of three known strong AMD genetic risk variants were constructed.

**Results:** SNP heritability is 55% +/- 13% (p= 9.87e-06) and the pedigree heritability is 49% +/- 18% (p= 3.06e-04). The sensitivity analyses revealed that the estimates were robust to changes in the inclusion of AMD variants as covariates.

**Conclusions:** These heritability estimates support existing twin and SNP-based AMD heritability estimates and corroborate the substantial involvement of genetics in AMD. Adjusting for known AMD variants revealed that additional genetic contribution exists, supporting a large polygenic effect in AMD.

## Introduction

Age-related Macular Degeneration (AMD) is a complex trait that can lead to central vision loss in its advanced stages^1^. It is a progressive disease with pathologic features often beginning as early as age 50 and visual disturbance onset commonly occurring in the sixth decade of life^2,3^. Major risk factors, in addition to age, include sex, smoking, and genetic contributions^4–6^. The development of AMD begins with retinal pigment epithelium (RPE) – adjacent deposits (such as drusen and subretinal drusenoid deposits, SDD), which are largely lipoprotein accumulations at the apical (SDD) or basal (drusen) side of the RPE, and are indicators of degrading outer retina function. As the deposits accumulate and the disease progresses, advanced complications such as the development of exudative macular neovascularization (neovascular AMD) or loss of the photoreceptors and RPE (geographic atrophy) can ensue^2^. These later stages of disease are when visual impairment occurs^1^. There are few treatments available, including micronutrient supplementation that can be incorporated in early stages to slow progression, anti-VEGF injections that are utilized in late-stage neovascular AMD to reduce vision loss, and complement targeting injections used to modestly slow progression in geographic atrophy cases^7–11^. AMD impacts millions of individuals in the U.S. with a prevalence rate of 12.3% in Non-Hispanic Whites (NHW)^12^ and is projected to affect 28 million individuals in North America by 2040^3^. As the population continues to age, the number of people at risk for developing AMD will continue to increase. Investigation into genetic contributions to disease becomes increasingly necessary to guide the field toward novel genetically supported drug targets, as they have higher success rates^13,14^.

There are numerous genetic risk loci associated with AMD^6^ many of which implicate the inflammatory and immune pathways, specifically the complement pathway. Notably, variants in the CFH and ARMS2/HTRA1 loci are associated with AMD^15–20^. Gorski and Grunin et al. (2025) reaffirm that the Y402H and R1210C variants in the CFH locus and the A69S variant in the ARMS2/HTRA1 locus confer substantial risks for AMD^6^. Individuals carrying the Y402H or A69S variants have 2-3 times the odds of developing AMD compared to non-carriers, while carriers of the rare variant R1210C are approximately 57 times more likely to develop AMD than non-carriers^6^.

The proportion of the AMD phenotype conferred by genetic effects is assessed through heritability estimations. In a small female twin study of individuals from the UK, heritability was estimated at 45%^21^. A larger USA based male twin study concluded heritability ranged from 46-71%^22^. The heritability across all AMD stages resulted in an estimate of 46%; including only intermediate/advanced AMD gave an estimate of 67%, and including only advanced AMD gave an estimate of 71%^22^. Similarly, SNP-based heritability from the International Age-Related Macular Degeneration Genetic Consortium in 2016 was estimated at 58%, when a 10% disease prevalence was assumed^23^.

While twin studies are commonly used for heritability estimates, they are particularly vulnerable to ascertainment biases often resulting in an overrepresentation of monozygotic, female, affected, and concordant twins. Some of these biases can lead to overestimation of genetic involvement in disease^24^. To update the heritability estimates for AMD and to identify the amount of genetic susceptibility that is left to discover, alternative pedigree-based designs, and newer relationship-informed genome-wide approaches are necessary. These expanded approaches will aid in clarifying AMD heritability.

The current study estimates both a SNP-based and a pedigree-based heritability for AMD using the same data from U.S. Amish communities. As a founder population from Europe, the Amish are a highly related community with higher average kinship coefficients between pairs of individuals compared to the surrounding communities.

Additionally, the Amish keep extensive genealogical documentation^25^. This study utilizes a single group of related individuals to estimate SNP-based and pedigree-based heritability. The goal of this study was to estimate the heritability of AMD in a sample of Ohio, Indiana, and Pennsylvania Amish community members to provide updated heritability predictions as well as to better understand the genetic risk of AMD in the Amish.

## Methods

### 2.1 Data collection

The Amish Eye Study enrolls individuals from Amish communities in and around Holmes County Ohio, Adams, LaGrange, and Elkhart Counties in Indiana, and Lancaster County Pennsylvania. The Amish are Anabaptists that immigrated to the United States from Switzerland and Germany in the 1700-1800s^26^. This founder population has high rates of endogamy with few new members joining from outside populations. The population expanded through high birth rates and settlements spread west in search of land^26^. Ascertainment approaches in Ohio and Indiana share a common method, identifying potential participants through Amish community directories, whereas Pennsylvania participants are enrolled upon visiting community health centers. Due to this difference in ascertainment, a larger number of closely related individuals are enrolled in Pennsylvania. The Anabaptist Genealogical Database (AGBD) documents family structure for all Amish, which allowed for the construction of a 14-generation pedigree containing all participants^27^. In addition to these pedigree data, once enrolled, participants in the study provide demographic data, family and individual medical history, a clinical eye exam with extensive imaging, and a blood sample. Informed consent was obtained for all participants, and all research methods followed approved Institutional Review Board protocols.

DNA was extracted from blood samples and genotyped on either the Illumina Multi-Ethnic Genotyping Array (MEGAex) with 3k additional custom SNPs or the Illumina Global Screening Array (GSA). Further details regarding genotyping, imputation and quality control have been previously published^28^ Quality control resulted in 2,042 samples across all the Amish and 338,575 SNPs.

Eye exams and imaging results were obtained by teams of optometrists and ophthalmologists at each site. Grading of all imaging was performed at the Doheny Image Reading and Research Lab. Color fundus photographs of the retina were examined for AMD related retinal abnormalities and classified according to the Beckman scale^29^. Participants with a grade of 0-2 for both eyes were defined as controls (no-AMD), participants with two grade 3 eyes or one grade 3 eye and one control eye were defined as intermediate AMD, and participants with two grade 4-5 eyes or one grade 4-5 eye and one intermediate eye were defined as advanced AMD. Cases (AMD) are an aggregate of intermediate AMD and advanced AMD. Participants with a defined phenotype, genotype information, reported sex, and aged 50 or older were eligible for participation in this study (n = 1156).

### 2.2 Heritability estimation

#### 2.2.1 Relationship determination

The SNP-based heritability (h^2^) model first required the use of the Kinship-based INference for Gwas (KING) software^30^ to identify genetic relationships among our participants. The robust function identifies relative pairs up to the second degree. The kinship coefficient of each pair was used as input into the LDAK Tetraher software^31^, specifically developed to perform h^2^estimation on related samples. Meanwhile, the pedigree-based heritability (h^2^) model utilizes pedigree information from the AGDB, allowing the construction of three generation (grandparental) pedigrees that match the individuals identified as first and second-degree relatives in KING to keep sample size and relationship patterns consistent. The pedigrees were input into the Sequential Oligogenic Linkage Analysis Routines (SOLAR) software^32^ for estimation of h^2^.

#### 2.2.2 Liability scale

Binary traits like AMD have an assumed underlying continuous liability where once a threshold is reached participants are considered affected^33^. The use of this underlying liability allows for inclusion of population prevalence so predictions can be compared across groups regardless of the proportion of cases^33^. LDAK TetraHer allows for the inclusion of population prevalence and directly estimated the heritability on the liability scale, whereas SOLAR estimates heritability on the observed scale and must be *post hoc* transformed to the liability scale. Equation 23 from Lee et al. 2011 is used to transform h^2^ and its corresponding standard error (SE) to the liability scale^33^.

#### 2.2.3 Models

Primary h^2^and h^2^models were built, adjusted for population prevalence and the covariates age, sex, and principal component 1, which was calculated using the relationship informed GENetic EStimation and Inference in Structured samples (GENESIS)^34^ PC-Air function to account for population structure within ancestry. AMD population prevalence of 12.3%^12^ was included in the h^2^model whereas the h^2^model was *post hoc* transformed. As sensitivity analyses, a model including three additional covariates, binary carrier statuses for A69S, Y402H, and R1210C was generated. Followed by several models including only one or two AMD variant carrier status covariates (Y402H, A69S, R1210C) in all possible combinations. Additionally, two models for a stratified analysis of cases split into intermediate and advanced stages were constructed.

#### 2.2.4 Primary model comparison

A nested model scheme and likelihood ratio test were used to compare the two primary models. The full model estimated heritability using both the GRM produced by KING and a kinship matrix produced by SOLAR and adjusted for age, sex, and PC1. The reduced model was derived from the full model by removing the kinship matrix. Using the REML function from the LDAK software heritability and the log-likelihood of each model was estimated. A likelihood ratio test comparing the two models followed.

## Results

### 3.1 Data characteristics

Of 1,156 eligible participants, KING identified 1,363 pairs of relatives composed of 525 individuals up to the second degree that were used in the h^2^model. For consistency, the same 525 participants were selected from the pedigree to be used in the h^2^model. Table 2 summarizes the age, sex, and carrier status of A69S, Y402H, and R1210C for the participants in the AMD and no-AMD groups. Age is significantly higher in individuals with AMD (69.85 years vs 64.08 years; p = <0.001). As expected, A69S and R1210C risk variant carriers were notably more common in the AMD groups (49.2% vs 41.4% and 3.8% vs 0.7% respectively) although the difference is not statistically significant. This difference remained insignificant when the data was restricted to advanced cases only. The minor allele frequency (MAF) and effect of the three noted AMD risk variants, Y402H, R1210C, and A69S on the general European population and the Amish 525 individuals stratified by ascertainment site (Pennsylvania vs Ohio/Indiana) are listed in table 1. The MAF of Y402H is similar across groups at 0.40 in the Pennsylvania group and 0.49 in the Ohio/Indiana group. R1210C is nonexistent in the Pennsylvania group and rare in the Ohio/Indiana group with a MAF of 0.02. Notably, for A69S, Pennsylvania had a MAF of 0.34 and Ohio/Indiana had a MAF of 0.16.

**Table 1:** Y402, R1210C, and A69S odds ratios (ORs) from Gorski et al. 2025, minor allele frequency (MAF) according to GnomAD, and the MAF in these SNPs in the Amish population stratified by enrollment site (Ohio/Indiana vs Pennsylvania).

| Variant | Locus | OR | EUR<br>MAF | Ohio/Indiana<br>Amish MAF | Pennsylvania<br>Amish<br>MAF |
| --- | --- | --- | --- | --- | --- |
| A69S | ARMS2/HTRA1 | 2.94 | 0.22 | 0.16 | 0.34 |
| Y402H | CFH | 2.40 | 0.62 | 0.49 | 0.40 |
| R1210C | CFH | 57.13 | 0.0002 | 0.02 | 0.00 |

**Table 2:** Age, sex, and carrier status for the A69S, Y402H, and R1210C variants in the 525 participants in the heritability models stratified by AMD and no-AMD groups.

|  | no-AMD | AMD | p-value |
| --- | --- | --- | --- |
| n = 525 | 285 | 240 |  |
| age (mean (SD)) | 64.08 (9.22) | 69.85 (9.55) | <0.001 |
| sex = M (%) | 124 (43.5) | 98 (40.8) | 0.596 |
| A69S = 1 (%) | 118 (41.4) | 118 (49.2) | 0.090 |
| Y402H = 1 (%) | 202 (70.9) | 163 (67.9) | 0.523 |
| R1210C = 1 (%) | 2 (0.7) | 9 (3.8) | 0.034 |

### 3.2 AMD Heritability

The primary h^2^and h^2^models resulted in an estimate of 55% +/- 13% (p= 9.87e-06) and 49% +/- 18% (p= 3.06e-04) respectively (Figure 1). The h^2^model including all three major variants produced an estimate of 59% +/- 13% (p = 3.14e-06) and the corresponding h^2^ model estimate is 49% +/- 18% (p = 3.04e-04) (Figure 2). Inclusion of the major AMD risk variants did not change the heritability estimates in the SNP or pedigree models. The likelihood ratio test comparing the models revealed a log-likelihood of −361 for the reduced model with the GRM and the full model with both the GRM and the kinship matrix produced a log-likelihood of −354. The likelihood ratio test produced a statistically significant p value of 1.03e-04 suggesting a statistically significant difference between models.

**Figure 1:**
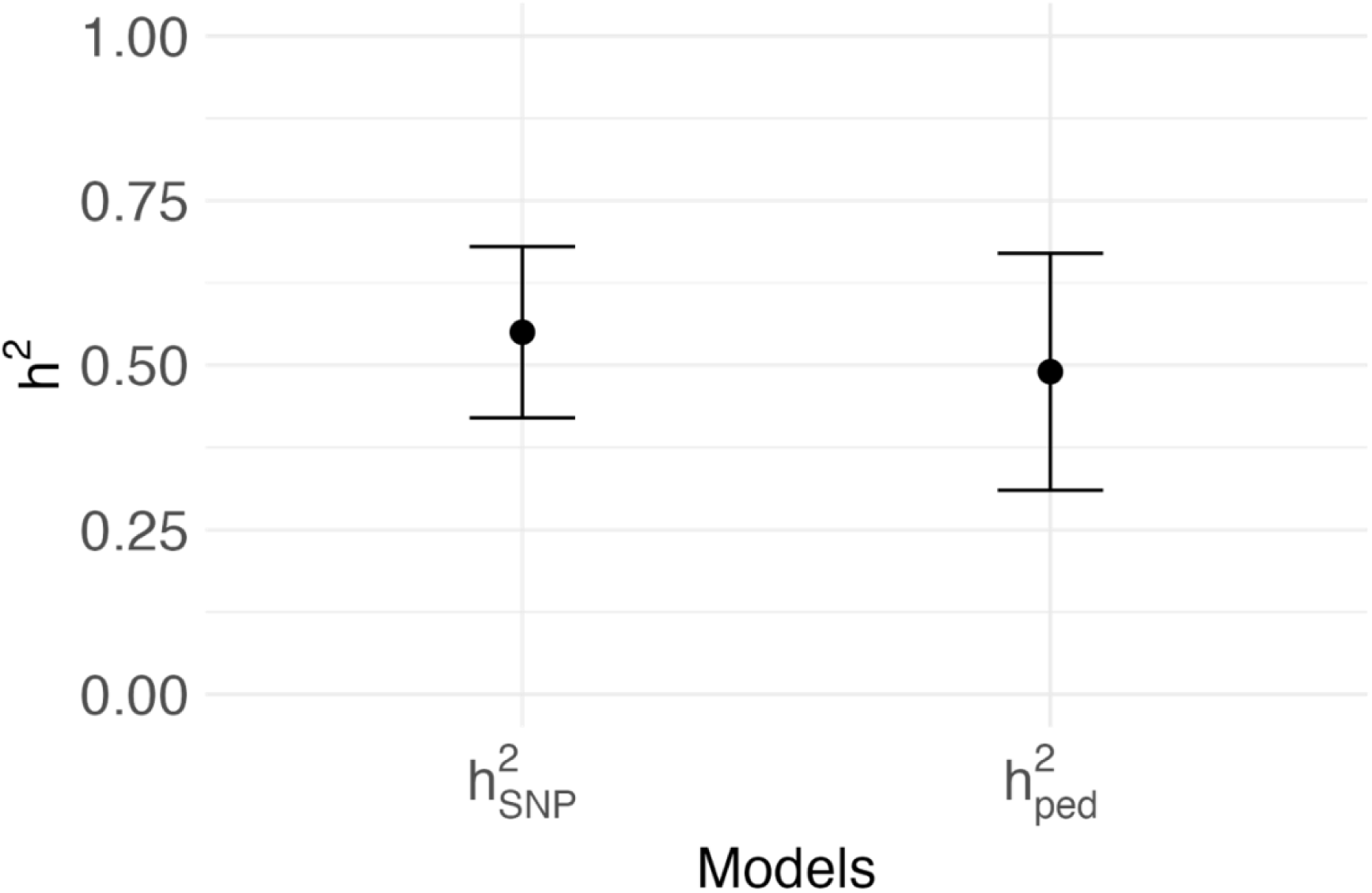
AMD SNP-based heritability estimate compared to pedigree-based heritability estimate both adjusted for age, sex, and PC1.

**Figure 2:**
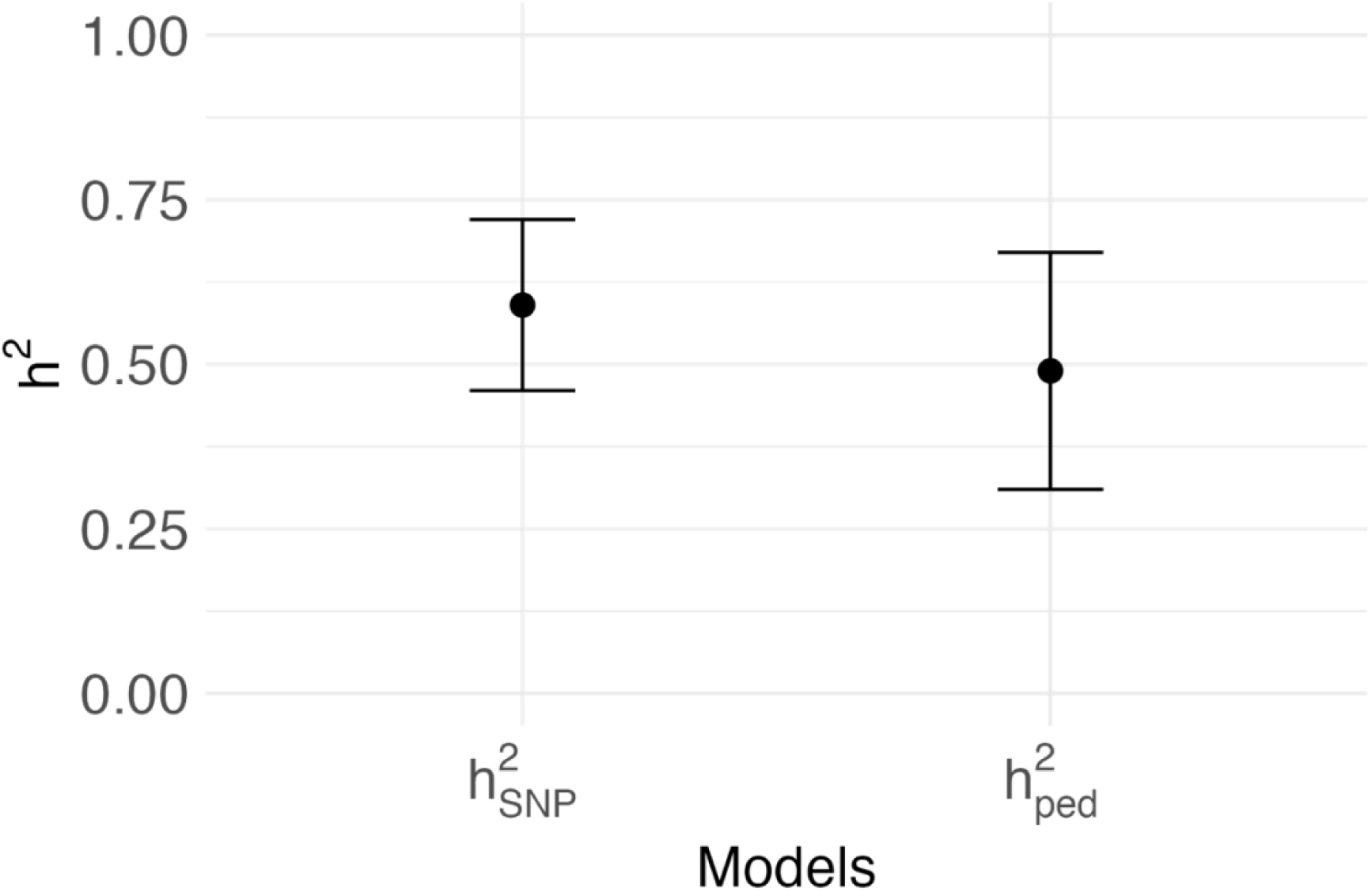
AMD SNP-based heritability estimate compared to pedigree-based heritability estimate both adjusted for age, sex, PC1, A69S, Y402H, R1210C binary carrier status.

The sensitivity models where known AMD variant carrier status covariates were included in all possible combinations resulted in SNP heritability point estimates that ranged from 54%-61% and the pedigree-based heritability results ranged from 48%-50%. These sensitivity analyses show that including known variants in the model did not substantially contribute to the model (Supplementary material Table 1 & Figure 1). The stratified analysis resulted in a h^2^estimate of 63% +/- 23% (p = 1.32e-02) and h^2^estimate of 62% +/- 32% (p=1.54e-02) for intermediate AMD and a h^2^estimate of 37% +/- 23% (p=1.09e-01) and h^2^estimate of 0.97% +/- 28% (p=4.17e-05) for advanced AMD (Supplementary material Figure 2). The small sample sizes and large standard errors for the intermediate AMD (AMD = 87 & no-AMD= 253) and advanced AMD (AMD = 129 & no-AMD= 272) estimates in the stratified analysis reduces confidence in these.

## Discussion

The SNP- and pedigree-based AMD heritability estimates in the Amish range from 49%-55% with highly overlapping standard errors. Both estimates are consistent with previous reports of heritability ranging from 45%-71%^21–23^ and provide additional support that there is a large genetic contribution to AMD.

The comparison of the full model and reduced model through a likelihood ratio test revealed that the reduced model has a good fit for predicting heritability using SNPs, however, the full model including the additional kinship matrix improves fit. This suggests that including the kinship matrix allows for the capture of additional phenotypic variance beyond what is captured by the GRM alone. This supports the utility of SNP-based heritability estimation methods and emphasized the value of pedigree-based methods as an alternative to twin studies.

The divergence of the SNP and pedigree-based heritability estimates is minimal; however, pedigree-based heritability is typically expected to be higher than SNP-based heritability, as it summarizes all transmitted effects as well as environmental contribution. One explanation is that the pedigree-based heritability may be deflated due to shared environment, as the Amish culture promotes dietary patterns that remain stable over time, living on multigenerational family land, and avoiding alcohol consumption and smoking, a well-known risk factor for AMD^5^. The stable lifestyle practiced by the Amish community makes this population reduces environmental variability between participants, lessening the potential environmental bias in pedigree-based heritability, thus limiting the potential impact on overall variance.

The A69S, Y402H, R1210C alleles were included as covariates in the analysis due to their large impact on risk of AMD^6^ (Table 1). The inclusion of covariates A69S, Y402H, and/or R1210C in various combinations did not significantly change the heritability estimates, suggesting that these large effect variants are not overshadowing other factors contributing to overall AMD heritability. A69S was of interest due to the large difference in MAF in the communities from the two ascertainment sites (Indiana/Ohio MAF=0.16; Pennsylvania MAF=0.34). We suspect this is due to a specific founder effect in the Lancaster Pennsylvania communities^26^. Although R1210C has a very large effect size, it is so rare in our population that it may not contribute to phenotypic variance. Additionally, Y402H has a MAF of 45% and is not associated with AMD in our sample (data not shown). Our results support that there are additional variants that in aggregate, largely contribute to the heritability of AMD, perhaps to a greater extent than the large effect variants, regardless of theoretically small effect sizes. These results highlight the genetic contribution to AMD beyond large effect variants and emphasize the importance of further investigation to identify additional small effect variants that collectively may be driving the heritability of this complex disease.

This study confirms that AMD is highly heritable, supporting previous twin analyses. The results also support the utility of SNP- and pedigree-based heritability estimation methods as effective and more accessible alternative methods to twin studies. The increased understanding of AMD genetics in the Amish presented here, through both SNP- and pedigree-based heritability estimates, encourages further investigation into the genetic contribution toward AMD. These results from a subgroup population are strikingly similar to heritability reports of the general Non-Hispanic White population^21–23^ suggesting that the Amish are representative of the NHW population and studying this population will be beneficial in identifying additional genetic factors relevant to the wider population.

Limitations of this study include a relatively small sample size of 525 participants. Increasing the sample size in the future will add confidence to the results. The different ascertainment strategies in Pennsylvania and Ohio/Indiana are also limiting due to the resulting inconsistent family structure between the sites. Specifically, in these analyses Pennsylvania participants had a similar number of participants yet an increased number of relationships compared to Ohio/Indiana, due to increased family enrollment. As we continue enrolling additional participants sample size will increase leading to more precise results.

## Data Availability

All data produced in the present study are available upon reasonable request to the authors.

## Acknowledgements

We thank the Amish community members for participating in this study. Thank you to the Wooster eye center and Great Lakes eye center. Research described in this publication is supported by National Institutes of Health and National Eye Institute under award numbers EY030614 and EY022310. N.C.M. is also supported by the National Institute of Health and National Institute on Aging through the Alzheimer’s Disease Translational Data Science Training Program (AG071474). D.S. is also supported by an unrestricted award from Research to Prevent Blindness and The Paul and Evanina Bell Mackall Foundation Trust. The content is solely the responsibility of the authors and does not necessarily represent the official views of the National Institutes of Health.

## Supplementary Material

**Supplementary Table 1:** Sensitivity analysis results. Heritability estimates for several models with removal of one or two AMD variant covariates.

| SNP adjusted | SNP h2 | p | ped h2 | p |
| --- | --- | --- | --- | --- |
| A69S | 0.54 (0.13) | 1.54E-05 | 0.50 (0.18) | 3.10e-04 |
| Y402H | 0.55 (0.13) | 1.27E-05 | 0.49 (0.17) | 2.87e-04 |
| R1210C | 0.60 (0.13) | 1.45E-06 | 0.48 (0.17) | 3.43e-04 |
| A69S & Y402H | 0.54 (0.13) | 2.08E-05 | 0.50 (0.18) | 2.95e-04 |
| A69S&R1210C | 0.59 (0.13) | 2.45E-06 | 0.49 (0.17) | 3.35e-04 |
| Y402H & R1210C | 0.61 (0.13) | 1.76E-06 | 0.49 (0.17) | 3.06e-04 |

**Supplementary Figure 1:**
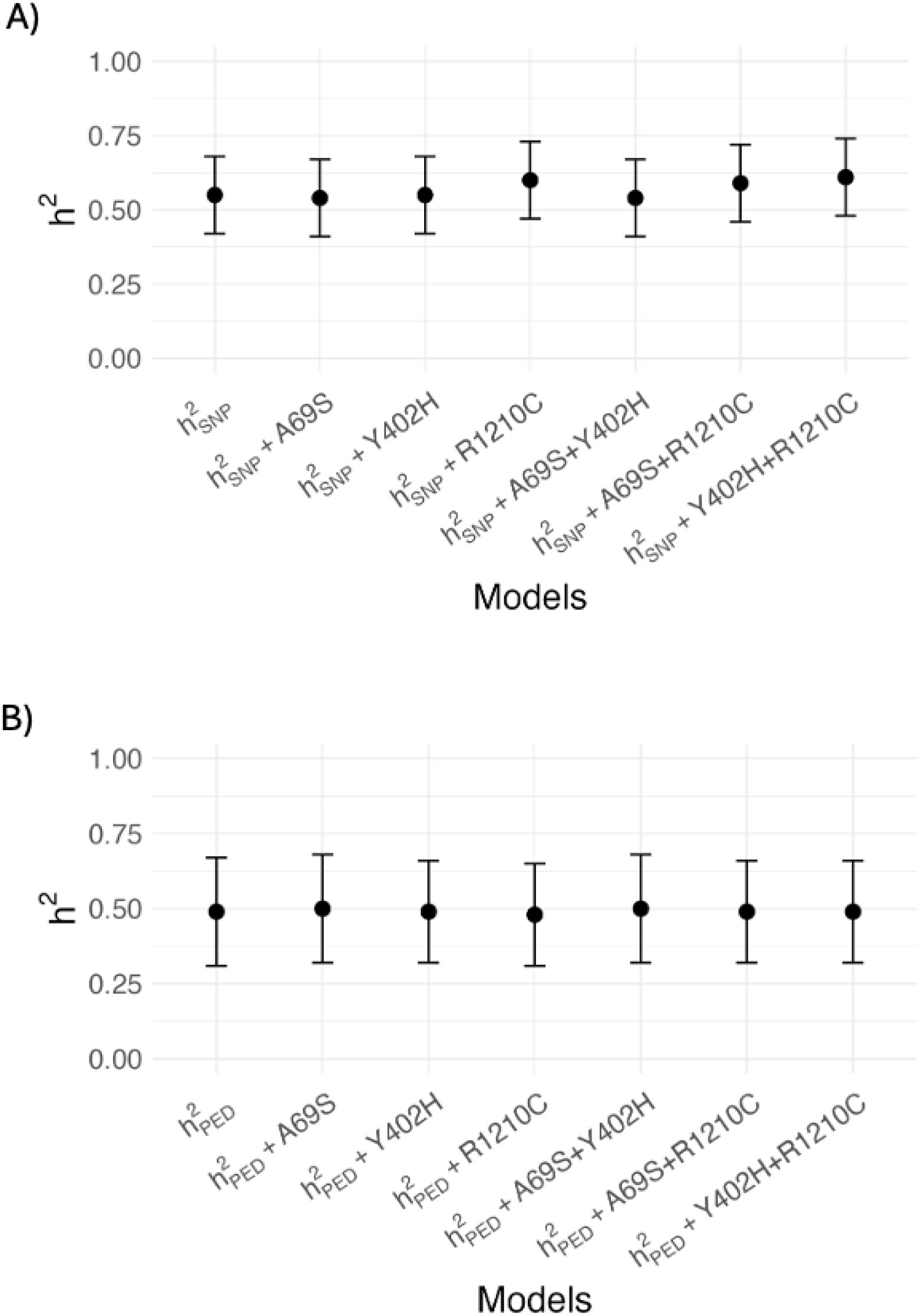
Comparison of sensitivity models adjusted for all three AMD variants (h^2^or h^2^), one, or two variants are compared. A) SNP-based heritability results, B) Pedigree-based heritability results.

**Supplementary Figure 2:**
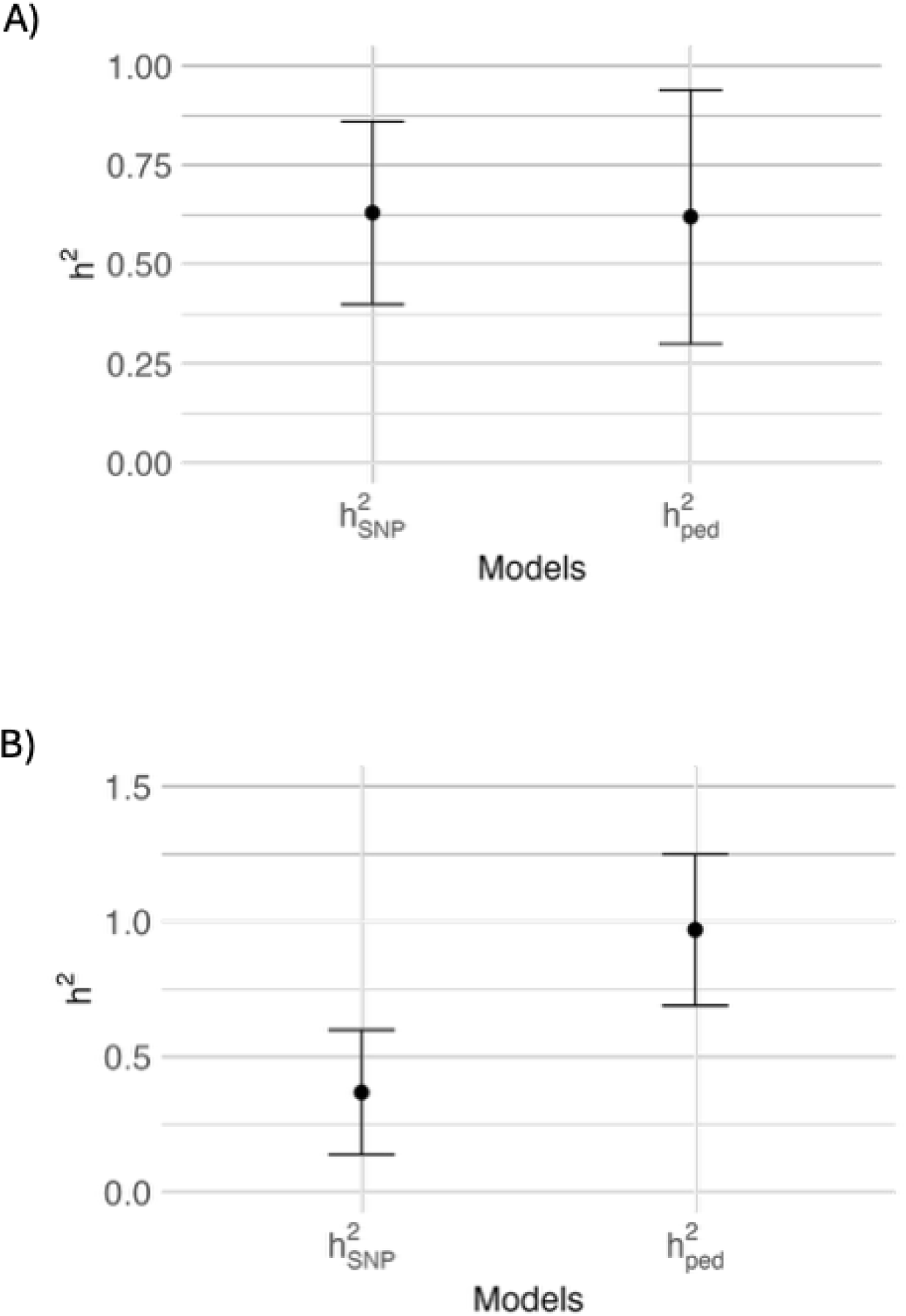
Comparison of AMD SNP- and pedigree-based heritability estimates both of which are adjusted for age, sex, PC1. A) Intermediate and B) Advanced.

## References

1. Fleckenstein M, Schmitz-Valckenberg S, Chakravarthy U. Age-Related Macular Degeneration: A Review. JAMA. 2024;331(2):147–157. doi:10.1001/jama.2023.26074

2. Fleckenstein M, Keenan TDL, Guymer RH, et al. Age-related macular degeneration. Nat Rev Dis Primers. 2021;7(31):1–25. doi:10.1038/s41572-021-00265-2

3. Wong WL, Su X, Li X, et al. Global prevalence of age-related macular degeneration and disease burden projection for 2020 and 2040: a systematic review and meta-analysis. Lancet Glob Health. 2014;2(2):e106–e116. doi:10.1016/S2214-109X(13)70145-1

4. Smith W, Mitchell P, Wang J. Gender, oestrogen, hormone replacement and age-related macular degeneration: Results from the Blue Mountains Eye Study. Australian and New Zealand Journal of Ophthalmology. 1997;25(Suppl 1):S13–S15. doi:10.1111/j.1442-9071.1997.tb01745.x

5. Thornton J, Edwards R, Mitchell P, Harrison RA, Buchan I, Kelly SP. Smoking and age-related macular degeneration: a review of association. Eye. 2005;19(9):935–944. doi:10.1038/sj.eye.6701978

6. Gorski M, Grunin M, Herold JM, et al. Diverse-Ancestry GWAS of Age-Related Macular Degeneration on 16,108 Examined Cases and 18,038 Controls. Invest Ophthalmol Vis Sci. 2025;66(13):51. doi:10.1167/iovs.66.13.51

7. Gragoudas ES, Anthony P. Adamis, Cunningham Jr ET, Matthew Feinsod, Guyer DR, for the VEGF Inhibition Study in Ocular Neovascularization Clinical Trial Group. Pegaptanib for Neovascular Age-Related Macular Degeneration. n engl j med. 2004;351(27):2805–2816. doi:10.1056/NEJMoa042760

8. Age-Related Eye Disease Study Research Group. A Randomized, Placebo-Controlled, Clinical Trial of High-Dose Supplementation With Vitamins C and E, Beta Carotene, and Zinc for Age-Related Macular Degeneration and Vision Loss: AREDS Report No. 8. Arch Ophthalmol. 2001;119(10):1417–1436. doi:10.1001/archopht.119.10.1417

9. Brown DM, Kaiser PK, Michels M, et al. Ranibizumab versus Verteporfin for Neovascular Age-Related Macular Degeneration. n engl j med. 2006;355(14):1432–1444. doi:10.1056/NEJMoa062655

10. Heier JS, Lad EM, Holz FG, et al. Pegcetacoplan for the treatment of geographic atrophy secondary to age-related macular degeneration (OAKS and DERBY): two multicentre, randomised, double-masked, sham-controlled, phase 3 trials. Lancet. 2023;402(10411):1434–1448. doi:10.1016/S0140-6736(23)01520-9

11. Khanani AM, Patel SS, Staurenghi G, et al. Efficacy and safety of avacincaptad pegol in patients with geographic atrophy (GATHER2): 12-month results from a randomised, double-masked, phase 3 trial. Lancet. 2023;402(10411):1449–1458. doi:10.1016/S0140-6736(23)01583-0

12. Rein DB, Wittenborn JS, Burke-Conte Z, et al. Prevalence of Age-Related Macular Degeneration in the US in 2019. JAMA Ophthalmol. 2022;140(12):1202–1208. doi:10.1001/jamaophthalmol.2022.4401

13. Minikel EV, Painter JL, Dong CC, Nelson MR. Refining the impact of genetic evidence on clinical success. Nature. 2024;629(8012):624–629. doi:10.1038/s41586-024-07316-0

14. Nelson MR, Tipney H, Painter JL, et al. The support of human genetic evidence for approved drug indications. Nat Genet. 2015;47(8):856–860. doi:10.1038/ng.3314

15. Haines JL, Hauser MA, Schmidt S, et al. Complement Factor H Variant Increases the Risk of Age-Related Macular Degeneration. Science. 2005;308(5720):419–421. doi:10.1126/science.1110359

16. Klein RJ, Zeiss C, Chew EY, et al. Complement Factor H Polymorphism in Age-Related Macular Degeneration. Science. 2005;308(5720):385–389. doi:10.1126/science.1109557

17. Jakobsdottir J, Conley YP, Weeks DE, Mah TS, Ferrell RE, Gorin MB. Susceptibility Genes for Age-Related Maculopathy on Chromosome 10q26. Am J Hum Genet. 2005;77(3):389–407. doi:10.1086/444437

18. Edwards AO, Ritter III R, Abel KJ, Manning A, Panhuysen C, Farrer LA. Complement Factor H Polymorphism and Age-Related Macular Degeneration. Science. 2005;308(5720):421–424. doi:10.1126/science.1110189

19. Rivera A, Fisher SA, Fritsche LG, et al. Hypothetical LOC387715 is a second major susceptibility gene for age-related macular degeneration, contributing independently of complement factor H to disease risk. Human Molecular Genetics. 2005;14(21):3227–3236. doi:10.1093/hmg/ddi353

20. Yang Z, Camp NJ, Sun H, et al. A Variant of the *HTRA1* Gene Increases Susceptibility to Age-Related Macular Degeneration. Science. 2006;314(5801):992–993. doi:10.1126/science.1133811

21. Hammond CJ, Webster AR, Snieder H, Bird AC, Gilbert CE, Spector TD. Genetic Influence on Early Age-related Maculopathy. Opthalmology. 2002;109(4):730–736. doi:10.1016/s0161-6420(01)01049-1

22. Seddon JM, Jennifer Cote, William F. Page, Steven H. Aggen, Michael C. Neale. The US Twin Study of Age-Related Macular Degeneration: Relative Roles of Genetic and Environmental Influences. Arch Ophthalmol. 2005;123(3):321–327. doi:10.1001/archopht.123.3.321

23. Fritsche LG, Igl W, Bailey JNC, et al. A large genome-wide association study of age-related macular degeneration highlights contributions of rare and common variants. Nat Genet. 2016;48(2):134–143. doi:10.1038/ng.3448

24. Hawkes CH. Twin studies in medicine--what do they tell us? Q J Med. 1997;90(5):311–321. doi:10.1093/qjmed/90.5.311

25. Strauss KA, Puffenberger EG. Genetics, Medicine, and the Plain People. Annu Rev Genom Hum Genet. 2009;10:513–536. doi:10.1146/annurev-genom-082908-150040

26. Crowley WK. OLD ORDER AMISH SETTLEMENT: DIFFUSION AND GROWTH. Annals of the Association of American Geographers. 1978;68(2):249–264. doi:10.1111/j.1467-8306.1978.tb01194.x

27. Main LR, Song YE, Lynn A, et al. Genetic analysis of cognitive preservation in the midwestern Amish reveals a novel locus on chromosome 2. Dement. 2024;20:7453–7464. doi:10.1002/alz.14045

28. Wang P, Song YE, Lynn A, et al. Heritability of Alzheimer’s disease–related plasma biomarkers in the Amish population. Alz & Dem Diag Ass & Dis Mo. 2026;18(2):e70388. doi:10.1002/dad2.70388

29. Ferris FL, Wilkinson CP, Bird A, et al. Clinical Classification of Age-related Macular Degeneration. Ophthalmology. 2013;120(4):844–851. doi:10.1016/j.ophtha.2012.10.036

30. Manichaikul A, Mychaleckyj JC, Rich SS, Daly K, Sale M, Chen WM. Robust relationship inference in genome-wide association studies. Bioinformatics. 2010;26(22):2867–2873. doi:10.1093/bioinformatics/btq559

31. Speed D, Evans DM. Estimating disease heritability from complex pedigrees allowing for ascertainment and covariates. The American Journal of Human Genetics. 2024;111(4):680–690. doi:10.1016/j.ajhg.2024.02.010

32. Almasy L, Blangero J. Multipoint Quantitative-Trait Linkage Analysis in General Pedigrees. Am J Hum Genet. 1998;62(5):1198–1211. doi:10.1086/301844

33. Lee SH, Wray NR, Goddard ME, Visscher PM. Estimating Missing Heritability for Disease from Genome-wide Association Studies. The American Journal of Human Genetics. 2011;88(3):294–305. doi:10.1016/j.ajhg.2011.02.002

34. Gogarten SM, Sofer T, Chen H, et al. Genetic association testing using the GENESIS R/Bioconductor package. Bioinformatics. 2019;35(24):5346–5348. doi:10.1093/bioinformatics/btz567

